# Evaluating Approaches for Constructing Polygenic Risk Scores for Prostate Cancer in Men of African and European Ancestry

**DOI:** 10.1101/2023.05.12.23289860

**Authors:** Burcu F. Darst, Jiayi Shen, Ravi K. Madduri, Alexis A. Rodriguez, Yukai Xiao, Xin Sheng, Edward J. Saunders, Tokhir Dadaev, Mark N. Brook, Thomas J. Hoffmann, Kenneth Muir, Peggy Wan, Loic Le Marchand, Lynne Wilkens, Ying Wang, Johanna Schleutker, Robert J. MacInnis, Cezary Cybulski, David E. Neal, Børge G. Nordestgaard, Sune F. Nielsen, Jyotsna Batra, Judith A. Clements, Australian Prostate Cancer BioResource, Henrik Grönberg, Nora Pashayan, Ruth C. Travis, Jong Y. Park, Demetrius Albanes, Stephanie Weinstein, Lorelei A. Mucci, David J. Hunter, Kathryn L. Penney, Catherine M. Tangen, Robert J. Hamilton, Marie-Élise Parent, Janet L. Stanford, Stella Koutros, Alicja Wolk, Karina D. Sørensen, William J. Blot, Edward D. Yeboah, James E. Mensah, Yong-Jie Lu, Daniel J. Schaid, Stephen N. Thibodeau, Catharine M. West, Christiane Maier, Adam S. Kibel, Géraldine Cancel-Tassin, Florence Menegaux, Esther M. John, Eli Marie Grindedal, Kay-Tee Khaw, Sue A. Ingles, Ana Vega, Barry S. Rosenstein, Manuel R. Teixeira, NC-LA PCaP Investigators, Manolis Kogevinas, Lisa Cannon-Albright, Chad Huff, Luc Multigner, Radka Kaneva, Robin J. Leach, Hermann Brenner, Ann W. Hsing, Rick A. Kittles, Adam B. Murphy, Christopher J. Logothetis, Susan L. Neuhausen, William B. Isaacs, Barbara Nemesure, Anselm J. Hennis, John Carpten, Hardev Pandha, Kim De Ruyck, Jianfeng Xu, Azad Razack, Soo-Hwang Teo, Canary PASS Investigators, Lisa F. Newcomb, Jay H. Fowke, Christine Neslund-Dudas, Benjamin A. Rybicki, Marija Gamulin, Nawaid Usmani, Frank Claessens, Manuela Gago-Dominguez, Jose Esteban Castelao, Paul A. Townsend, Dana C. Crawford, Gyorgy Petrovics, Graham Casey, Monique J. Roobol, Jennifer F. Hu, Sonja I. Berndt, Stephen K. Van Den Eeden, Douglas F. Easton, Stephen J. Chanock, Michael B. Cook, Fredrik Wiklund, John S. Witte, Rosalind A. Eeles, Zsofia Kote-Jarai, Stephen Watya, John M. Gaziano, Amy C. Justice, David V. Conti, Christopher A. Haiman

**Author notes:** Corresponding Author Burcu F. Darst, 1100 Fairview Ave N Seattle, WA 98109, 1-206-667-1036. Co-first authorship (Burcu F. Darst, Jiayi Shen).

## Abstract

Genome-wide polygenic risk scores (GW-PRS) have been reported to have better predictive ability than PRS based on genome-wide significance thresholds across numerous traits. We compared the predictive ability of several GW-PRS approaches to a recently developed PRS of 269 established prostate cancer risk variants from multi-ancestry GWAS and fine-mapping studies (PRS_269_). GW-PRS models were trained using a large and diverse prostate cancer GWAS of 107,247 cases and 127,006 controls used to develop the multi-ancestry PRS_269_. Resulting models were independently tested in 1,586 cases and 1,047 controls of African ancestry from the California/Uganda Study and 8,046 cases and 191,825 controls of European ancestry from the UK Biobank and further validated in 13,643 cases and 210,214 controls of European ancestry and 6,353 cases and 53,362 controls of African ancestry from the Million Veteran Program. In the testing data, the best performing GW-PRS approach had AUCs of 0.656 (95% CI=0.635-0.677) in African and 0.844 (95% CI=0.840-0.848) in European ancestry men and corresponding prostate cancer OR of 1.83 (95% CI=1.67-2.00) and 2.19 (95% CI=2.14-2.25), respectively, for each SD unit increase in the GW-PRS. However, compared to the GW-PRS, in African and European ancestry men, the PRS_269_ had larger or similar AUCs (AUC=0.679, 95% CI=0.659-0.700 and AUC=0.845, 95% CI=0.841-0.849, respectively) and comparable prostate cancer OR (OR=2.05, 95% CI=1.87-2.26 and OR=2.21, 95% CI=2.16-2.26, respectively). Findings were similar in the validation data. This investigation suggests that current GW-PRS approaches may not improve the ability to predict prostate cancer risk compared to the multi-ancestry PRS_269_ constructed with fine-mapping.

## Main Text

Prostate cancer is the second leading cause of cancer deaths among men in the US, with incidence rates being highest in men of African ancestry^1,2^. Earlier identification of men with increased risk of prostate cancer across diverse populations has the potential to reduce the stark health disparities of this disease. We recently performed a large and diverse genome-wide association (GWAS) of prostate cancer in men from African, European, East Asian, and Hispanic populations^3^. By performing ancestry-specific and multi-ancestry GWAS and fine-mapping analyses, this investigation revealed 269 GWAS-defined prostate cancer risk variants used to develop a multi-ancestry polygenic risk score (PRS_269_). The PRS_269_ was highly predictive of prostate cancer risk across populations^3^ and has since been validated in additional independent multi-ancestry studies^4^. However, genome-wide PRS (GW-PRS) approaches, which include variants across the genome that do not reach genome-wide statistical significance thresholds, have been reported to have better predictive performance than standard pruning and thresholding PRS of known variants across numerous complex traits, including schizophrenia, coronary artery disease, atrial fibrillation, type 2 diabetes, inflammatory bowel disease, breast cancer, and colorectal cancer^5-9^.

In this investigation, we compared the predictive ability of GW-PRS for prostate cancer to the multi-ancestry PRS_269_ of established prostate cancer risk variants. GW-PRS models were trained using summary statistics from the studies used to construct the multi-ancestry PRS_269_, which included 107,247 cases and 127,006 controls from European (85,554 cases and 91,972 controls), African (10,368 cases and 10,986 controls), East Asian (8,611 cases and 18,809 controls), and Hispanic (2,714 cases and 5,239 controls) populations^3^. Three recent GW-PRS approaches were evaluated: LDpred2^10^, PRS-CSx^11^, and EB-PRS^12^, using the 1.1 million HapMap3 panel variants^13^ recommended by these approaches, which included 44 of the 269 variants, with all other autosomal prostate cancer risk variants being within 800 kb and correlated with a median r=0.99 (ranging from 0.31-1.00 in any given population) of at least one of the 1.1 million HapMap3 variants (**Supplemental Methods** and **Tables S1-S2**). For comparison, each model was trained using previously estimated multi-ancestry weights and population-specific weights from the GWAS summary statistics^3^. GW-PRS models were tested in African ancestry men from the California/Uganda Study (CA/UG Study; 1,586 cases and 1,047 controls) and European ancestry men from the UK Biobank (8,046 cases and 191,825 controls; **Supplemental Methods**). Additional validation was performed in 6,353 cases and 53,362 controls of African ancestry and 13,643 cases and 210,214 controls of European ancestry from the Million Veteran Program^14^ (MVP; **Supplemental Methods**).

In the CA/UG and UK Biobank testing datasets, the best performing GW-PRS approach was PRS-CSx with multi-ancestry weights, with an area under the curve (AUC) of 0.656 (95% CI=0.635-0.677) in African and 0.844 (95% CI=0.840-0.848) in European ancestry men (**Supplemental Methods, Figure 1**, and **Table S3**). Each SD unit increase in PRS was associated with 1.83-fold higher odds of prostate cancer (95% CI=1.67-2.00) in men of African ancestry and 2.19-fold higher odds of prostate cancer (95% CI=2.14-2.25) in men of European ancestry (**Supplemental Methods, Figure 1**, and **Table S4**). However, compared to PRS-CSx, the PRS_269_ had higher or nearly identical AUCs in both African (0.679, 95% CI=0.659-0.700) and European (0.845, 95% CI=0.841-0.849) ancestry men, and the PRS_269_ was associated with 2.05-fold higher odds (95% CI=1.87-2.26) and 2.21-fold higher odds (95% CI=2.16-2.26) of prostate cancer in African and European ancestry men, respectively (**Figure 1, Table S3**, and **Table S4**). Findings were consistent when investigating extreme PRS distributions, with similar prostate cancer OR observed for the PRS_269_ and the best performing GW-PRS (PRS-CSx) when comparing African and European ancestry men in the highest PRS decile (90-100%) to those in the average 40-60% PRS category (**Supplemental Methods, Figure S1**, and **Table S5**).

**Figure 1.**
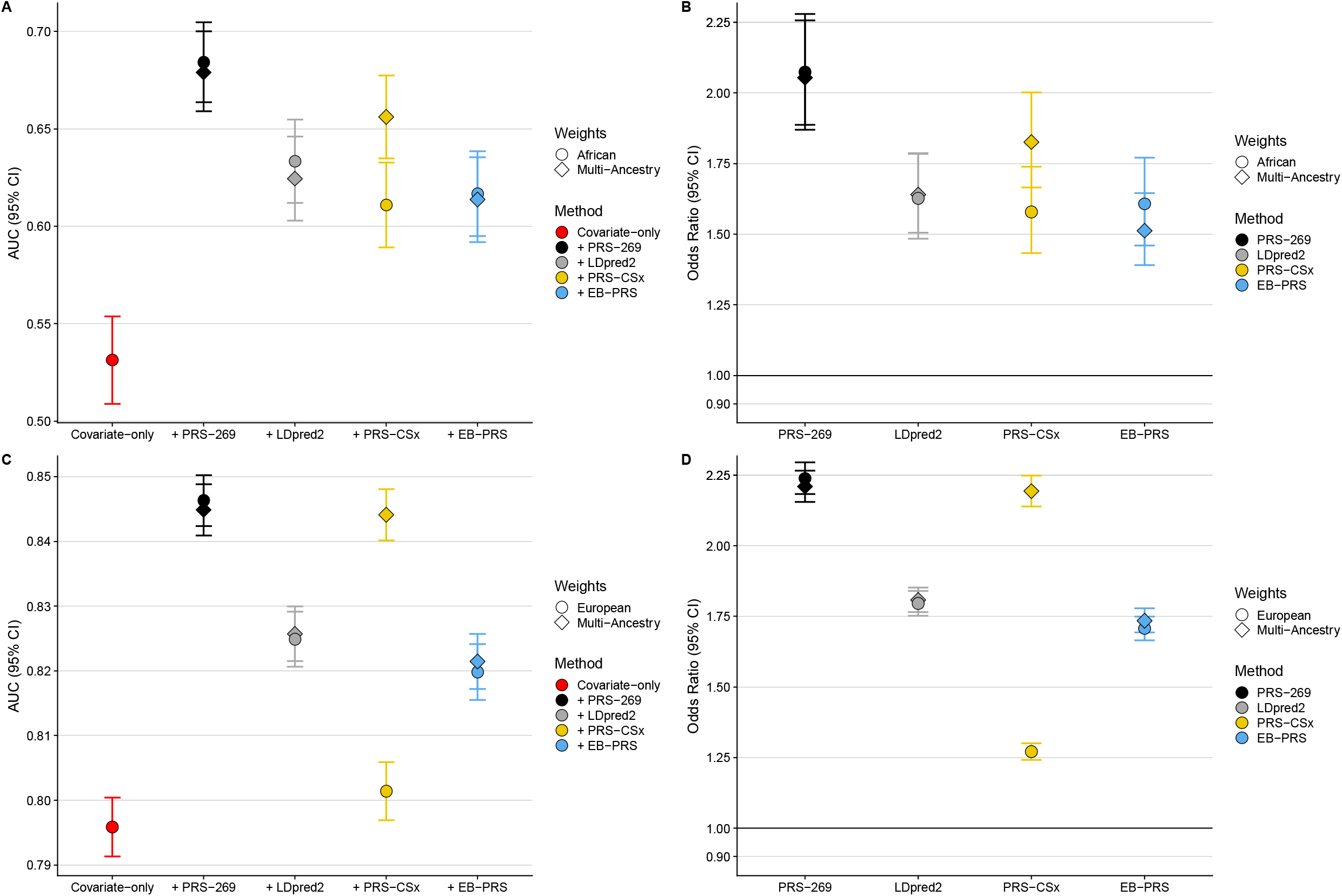
Comparison of PRS performance in the CA UG Study and the UK Biobank testing data. PRS performance is evaluated using area under the curve (AUC) estimated in men of A) African and C) European ancestry and OR of prostate cancer for each SD increase in PRS in men of B) African and D) European ancestry.

Similarly, in the validation MVP study, the best performing GW-PRS approach was PRS-CSx with multi-ancestry weights; however, the PRS_269_ performed either better or similarly with regards to AUC (AUC=0.656 [95% CI=0.649-0.663] versus AUC=0.624 [95% CI=0.617-0.632] in African ancestry men; AUC=0.694 [95% CI=0.690-0.699] versus AUC=0.692 [95% CI=0.687-0.696] in European ancestry men; **Figure 2** and **Table S3**). Likewise, the PRS_269_ was associated with prostate cancer OR that were comparable or larger than OR estimated with PRS-CSx (OR=1.77, 95% CI=1.72-1.82 versus OR=1.59, 95% CI=1.54-1.63 in African ancestry men; OR=1.99, 95% CI=1.95-2.02 versus OR=1.97, 95% CI=1.93-2.01 in European ancestry men; **Figure 2** and **Table S4**). OR calculated for African and European men in the top PRS decile were also comparable across the PRS_269_ and PRS-CSx (**Figure S2** and **Table S5**). In the testing and validation datasets, model performance was similar for both PRS_269_ and GW-PRS approaches when using either multi-ancestry or population-specific weights (**Figures 1-2, Figures S1-S2**, and **Tables S3-S5**).

**Figure 2.**
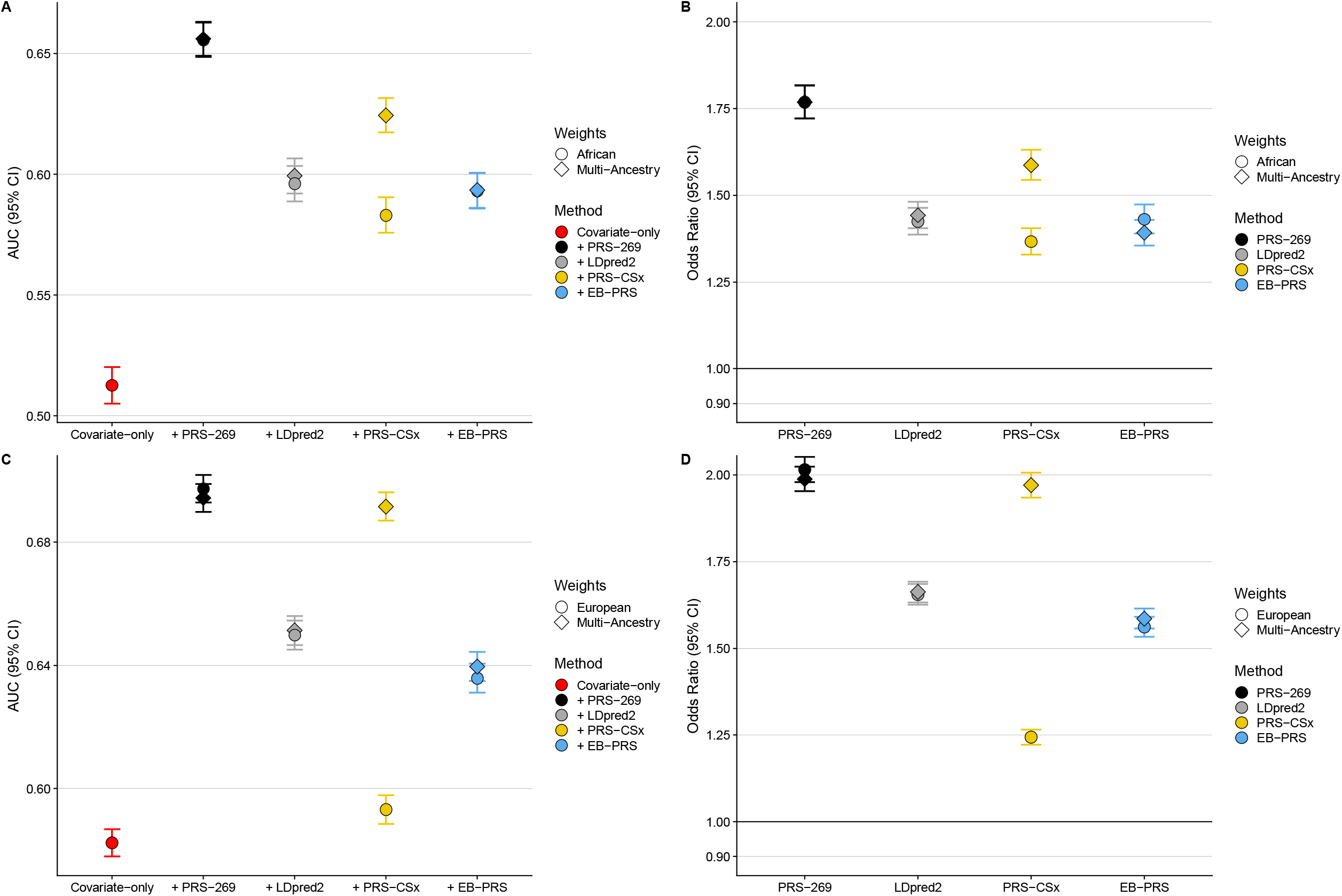
Comparison of PRS performance in the MVP validation data. PRS performance is evaluated using area under the curve (AUC) estimated in men of A) African and C) European ancestry and OR of prostate cancer for each SD increase in PRS in men of B) African and D) European ancestry.

Findings from this investigation suggest that current GW-PRS approaches do not outperform the multi-ancestry PRS_269_ for overall prostate cancer risk prediction. For several other disease examples, GW-PRS have been shown to perform better than PRS of known variants^5-9^; however, these PRS are typically constructed from a pruning and thresholding approach within European ancestry individuals rather than a fine-mapping approach across diverse populations. As such, the performance observed for our prostate cancer PRS_269_ may be due to identifying GWAS risk variants from a multi-ancestry GWAS and fine-mapping study, along with the use of the same multi-ancestry GWAS to construct the GW-PRS^3^. It is also possible that the unique genetic architecture of prostate cancer contributes to the high performance of the PRS_269_ across populations, as prostate cancer is one of the most heritable cancers^15,16^ and has been estimated to display a greater distribution of variants with larger effect sizes than other cancers with similar GWAS sample sizes^9^.

We have previously shown that GW-PRS including variants with weaker statistical evidence of association in both European and African ancestry men (based on lenient P-value thresholds down to 1.0×10^−5^) resulted in lower PRS performance^3^. Likewise, it was recently reported that a GW-PRS constructing from 1 million variants most strongly associated with prostate cancer risk led to comparable results as a GW-PRS based on HapMap3 variants^17^, further suggesting that GW-PRS approaches may not be improved by selecting a large number of variants weakly associated with prostate cancer risk. Last, a European-ancestry derived PRS of 110 established literature-curated prostate cancer risk variants was previously found to perform better than a GW-PRS in addition to a standard pruning and thresholding PRS^18^. These findings in conjunction with the present study suggest that the current multi-ancestry and fine-mapped PRS_269_ is optimal, which has important clinical implications. While genotyping a few hundred versus millions of variants to construct PRS is currently logistically easier and more cost-effective, genome-wide genotyping may be optimal in the future to enable the evaluation of PRS across many traits. However, our findings do not imply that the multi-ancestry PRS_269_ has reached optimal performance; increasing the sample size of non-European ancestry men in the discovery GWAS, particularly African ancestry men, where we and others have observed that the PRS has lower performance than in other populations^3,4^, will be important to improve genetic risk prediction of prostate cancer. The multi-ancestry PRS_269_ is an effective risk stratification tool for prostate cancer, and its clinical utility in screening and early detection warrants investigation.

## Data Availability

The summary statistics used in this investigation are available through dbGaP under accession code phs001120. 1000 Genomes Project data can be found online: https://www.internationalgenome.org/. UK Biobank data is available to researchers with approved applications.

## Acknowledgements

This work was supported by the National Cancer Institute at the National Institutes of Health grant (grant numbers U19 CA214253 to C.A.H., R01 CA257328 to C.A.H., U19 CA148537 to C.A.H., R01 CA165862 to C.A.H., and R00 CA246063 to B.F.D.), the Prostate Cancer Foundation (grants 21YOUN11 to B.F.D. and 20CHAS03 to C.A.H.), an award from the Andy Hill Cancer Research Endowment Distinguished Researchers Program (B.F.D.), a Fred Hutch/University of Washington SPORE Career Enhancement Program award (BFD), and the Million Veteran Program-MVP017. This research has been conducted using the UK Biobank Resource under application number 42195. This research is based on data from the Million Veteran Program, Office of Research and Development and the Veterans Health Administration. This publication does not represent the views of the Department of Veteran Affairs or the United States Government.

## Author Contributions

Study conception/design: BFD, DVC, CAH; Provided data: RKM, XS, EJS, TD, MNB, TJH, KM, PW, LLM, LW, YW, LS, RJM, CC, DEN, BGN, SFN, JB, JAC, APCB, HG, NP, RCT, JYP, DA, SW, LAM, DJH, KLP, CMT, RJH, MP, JLS, SK, AW, KDS, WJB, EDY, JEM, YL, SNT, CMW, CM, ASK, GC, FM, EMJ, EMG, KK, SAI, AV, BSB, MRT, NCPI, MK, LC, CH, LM, RK, RJL, HB, AWH, RAK, ABM, CJL, SLN, WBI, BN, AJH, JC, HP, KDR, JX, AR, ST, CPI, LFN, JF, CN, BAR, MG, NU, FC, MG, JEC, PAT, DCC, GP, GC, MFR, JFH, SIB, SKVDE, DFE, SJC, MBC, FW, JSW, RAE, AK, SW, JMG, ACJ, DVC, CAH; Data analysis: BFD, JS, YX, AAR; Bioinformatics support: XS, RKM; Interpretation of data: BFD, DVC, CAH; Drafted the manuscript: BFD; Contributed to manuscript revisions: BFD, JS, XS, EJS, EMJ, DJS, NP, DVC, CAH; All authors approved of the final manuscript.

