## Supplemental Materials for "Evaluating Approaches for Constructing Polygenic Risk Scores for Prostate Cancer in Men of African and European Ancestry"

Men of African and European Ancestry

Supplemental Material

*Participants and Genotype Data*

We used summary statistics from the largest and most diverse published prostate cancer genome-wide association study (GWAS) to date to train the genome-wide polygenic risk scores (GW-PRS) models. These summary statistics were based on 107,247 cases and 127,006 controls from European (85,554 cases and 91,972 controls), African (10,368 cases and 10,986 controls), East Asian (8,611 cases and 18,809 controls), and Hispanic (2,714 cases and 5,239 controls) populations^1^. The GW-PRS models were tested in African ancestry men from the California Uganda Study (CA/UG Study; 1,586 cases and 1,047 controls) and European ancestry men from the UK Biobank (8,046 cases and 191,825 controls). Participants from the CA/UG Study were genotyped using the Illumina H3 Africa array and imputed using Phase 3 of the 1000 Genomes Project^2^. Participants from the UK Biobank were genotyped using the Affymetrix UK Biobank Axiom Array and the Affymetrix UK BiLEVE Axiom Array and imputed using the Haplotype Reference Consortium (HRC), UK10K, and 1000 Genomes Project panels^3^. Quality control and study sample details have been previously described in detail^1^.

Additional validation was performed using men from the Million Veteran Program (MVP). Participants were men aged 40 years or older recruited between 2011 and 2017 and included 19,996 cases and 263,576 controls, of which 13,643 cases and 210,214 controls were of European ancestry and 6,353 cases and 53,362 controls were of African ancestry, as determined using the harmonized ancestry and race/ethnicity (HARE) algorithm that uses both genetically inferred ancestry and self-identified race/ethnicity to assign population categories^4^. Cases were men with a confirmed prostate cancer diagnosis from the VA’s Central Cancer Registry, while controls were men with no evidence of prostate cancer at their last prostate specific antigen (PSA) test. Participants were genotyped on a customized Affymetrix Axiom Biobank Array augmented to include variants specific to the VA populations, particularly African American and Hispanic populations^5^, and GWAS data were imputed using Phase 3 of the 1000 Genomes Project^2^. Details regarding MVP participant recruitment and genomic quality control have been previously described^5^.

*Polygenic Risk Scores*

We evaluated three GW-PRS approaches: 1) LDpred2^6^, an extension of the Bayesian LDpred method^7^ that modifies variant effect estimates based on LD between variants; 2) PRS-CSx^8^, an extension of the Bayesian PRS-CS approach^9^ that integrates GWAS summary statistics from multiple ancestry groups to improve cross-population polygenic modeling; and 3) EB-PRS^10^, an Empirical Bayes method that leverages effect size distributions across variants to improve prediction accuracy. LDpred2 was evaluated using the “auto”, “grid”, and infinitesimal (“inf”) options, and the best performing method (“grid”) was the focus of primary results reported in the manuscript, with all results reported in the supplemental material. PRS-CSx was evaluated with the meta=TRUE option in order to generate a single multi-ancestry PRS applied to all populations, with population-specific weights generated from the same model used to for population-specific PRS. GW-PRS models included the 1.1 million HapMap3 panel variants^11^, as recommended by these approaches. The HapMap3 variants were selected based on stringent quality control metrics and are limited to autosomal SNPs with rsIDs that map to a unique genomic location that had a missingness of <5%, Hardy-Weinberg equilibrium >1.0x10^-6^, and <3 Mendelian errors within the diverse HapMap3 populations^11^. As such, these variants are typically imputed with high quality and offer good coverage of the whole genome. Of our 269 PRS variants, 44 autosomal variants were included among the HapMap3 variants, while all other 217 autosomal variants were within 800 kb and correlated with a median r=0.99 (ranging from 0.31-1.00 in any given population) of at least one of the 1.1 million HapMap3 variants (**Table S1**).

LDpred2 and PRS-CSx both require LD reference panels, which were based on participants from the 1000 Genomes Project^2^ (**Table S2**). For each approach, GW-PRS were trained using both multi-ancestry marginal weights and population-specific marginal weights (European ancestry weights for the UK Biobank and European ancestry men from MVP and African ancestry weights for the CA UG Study and African ancestry men from MVP; **Table S2**).

For comparison, genome-wide significant PRS were constructed from 269 previously reported variants identified in a large and diverse GWAS and fine-mapping investigation (PRS_269_)^1^. For each testing and validation sample, the PRS_269_ was constructed using previously reported multi-ancestry conditional weights, calculated by meta-analyzing ancestry-specific conditional effects using a Z-score weighted fixed-effects meta-analysis, and ancestry-specific conditional weights (European ancestry weights for the UK Biobank and European ancestry men from MVP and African ancestry weights for the CA UG Study and African ancestry men from MVP)^1^. In the UK Biobank and CA UG Study, 267 of the 269 variants were present for PRS construction, while all 269 were present in MVP.

*Statistical Analyses*

Each PRS was evaluated as a continuous variable standardized by the mean and standard deviation within each population. PRS were also evaluated using an indicator variable splitting the PRS into the following categories: [0-10%], (10-20%], (20-30%], (30-40%], (40-60%], (60-70%], (70-80%], (80-90%], and (90-100%]. An additional analysis was performed splitting the top decile into two categories to evaluate the highest PRS percentile: (90-99%], (99-100%]. PRS category cutoffs were determined using PRS distributions in controls. Prostate cancer odds ratios for each PRS category were calculated using logistic regression models including covariates for age and principal components of ancestry, using the 40-60% PRS category as the reference for categorical PRS analyses. A P-value<0.05 was considered statistically significant.

Area under the curve (AUC) was used to evaluate the discriminative ability of the PRS. AUCs were calculated using logistic regression for 1) a base model with age and principal components of ancestry and 2) a full model with age, principal components, and the continuous PRS. All analyses were performed separately in each self-reported ancestral population.

Table S2. Genome-wide PRS models constructed. Each row represents a unique model.

| **Method** | **LD Reference** | **Input Weights** | **Output Weights** |
| --- | --- | --- | --- |
| LDpred2 | 1KGP EUR | Multi-ancestry | Multi-ancestry |
|  | 1KGP EUR | European | European |
|  | 1KGP AFR | Multi-ancestry | Multi-ancestry |
|  | 1KGP AFR | African | African |
| PRS-CSx | 1KGP EUR, 1KGP AFR, 1KGP EAS | European, African, East Asian, Hispanic | Multi-ancestry, European, African, East Asian, Hispanic |
| EB-PRS | -- | Multi-ancestry | Multi-ancestry |
|  | -- | European | European |
|  | -- | African | African |

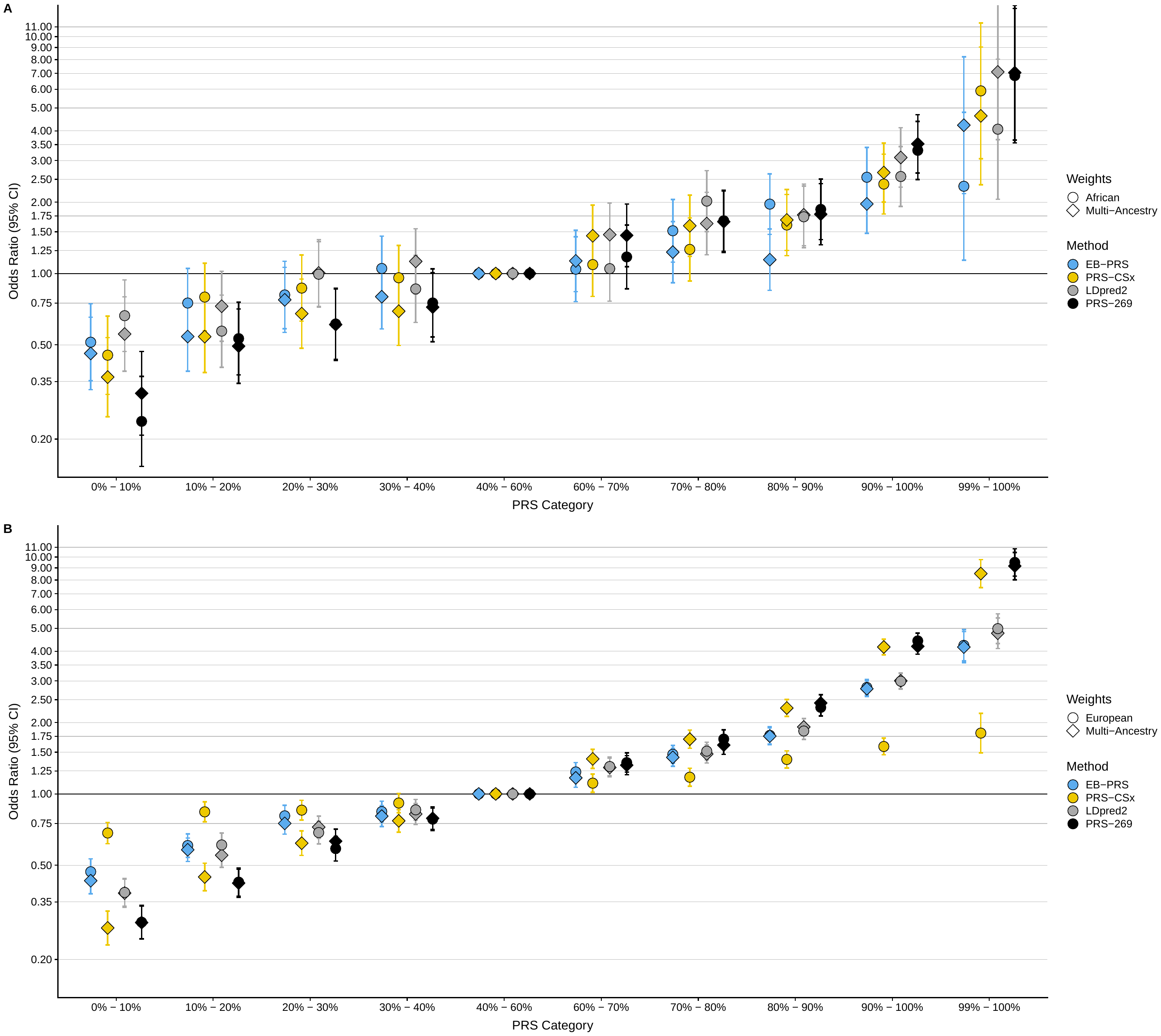

Figure S1. Comparison of prostate cancer OR for each PRS category in the CA UG Study and the UK Biobank testing data. Prostate cancer OR for each PRS category are relative to the 40-60% PRS category in men of A) African and B) European ancestry.

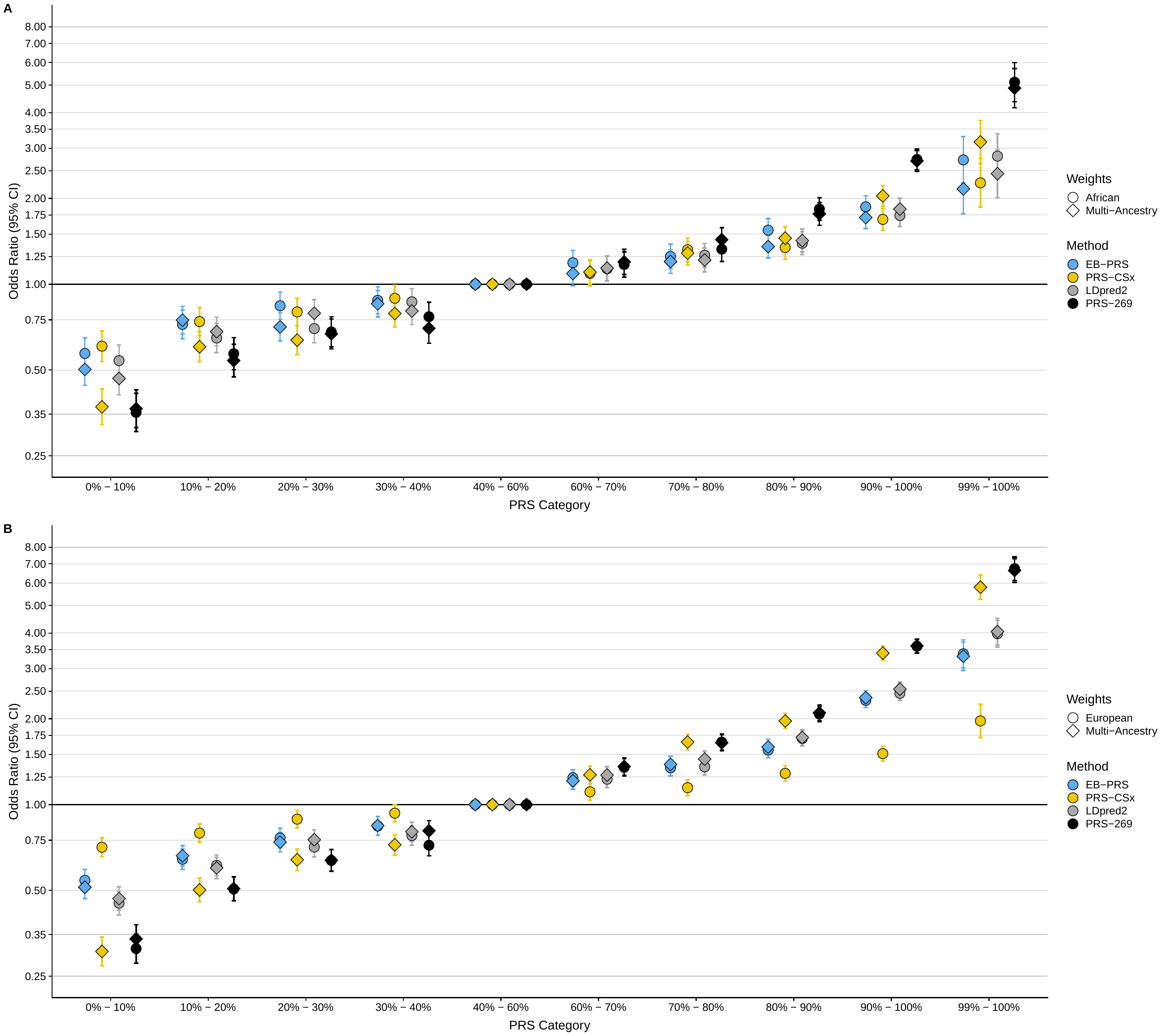

Figure S2. Comparison of prostate cancer OR for each PRS category in the MVP validation data. Prostate cancer OR for each PRS category are relative to the 40-60% PRS category in men of A) African and B) European ancestry.

Supplemental Note

The authors wish to pay tribute to Brian Henderson, who was a driving force behind this research, for his vision and leadership, and sadly passed away before seeing its fruition.

CRUK and PRACTICAL consortium

This work was supported by the Canadian Institutes of Health Research, European Commission's Seventh Framework Programme grant agreement n° 223175 (HEALTH- F2-2009-223175), Cancer Research UK Grants C5047/A7357, C1287/A10118, C1287/A16563, C5047/A3354, C5047/A10692, C16913/A6135, and The National Institute of Health (NIH) Cancer Post-Cancer GWAS initiative grant: No. 1 U19 CA 148537-01 (the GAME-ON initiative). This work was supported by the National Institute of General Medical Sciences at the NIH (grant number R35 GM140487 to D.J.S.).

We would also like to thank the following for funding support: The Institute of Cancer Research and The Everyman Campaign, The Prostate Cancer Research Foundation, Prostate Research Campaign UK (now Prostate Action), The Orchid Cancer Appeal, The National Cancer Research Network UK, The National Cancer Research Institute (NCRI) UK. We are grateful for support of NIHR funding to the NIHR Biomedical Research Centre at The Institute of Cancer Research and The Royal Marsden NHS Foundation Trust.

The Prostate Cancer Program of Cancer Council Victoria also acknowledge grant support from The National Health and Medical Research Council, Australia (126402, 209057, 251533, 396414, 450104, 504700, 504702, 504715, 623204, 940394, 614296,), VicHealth, Cancer Council Victoria, The Prostate Cancer Foundation of Australia, The Whitten Foundation, PricewaterhouseCoopers, and Tattersall’s.

Genotyping of the OncoArray was funded by the US National Institutes of Health (NIH) [U19 CA 148537 for ELucidating Loci Involved in Prostate cancer SuscEptibility (ELLIPSE) project and X01HG007492 to the Center for Inherited Disease Research (CIDR) under contract number HHSN268201200008I].

This study would not have been possible without the contributions of the following: Coordination team, bioinformatician and genotyping centers (CCGE, Cambridge).

Funding for the iCOGS infrastructure came from: the European Community's Seventh Framework Programme under grant agreement n° 223175 (HEALTH-F2-2009-223175) (COGS), Cancer Research UK (C1287/A10118, C1287/A 10710, C12292/A11174, C1281/A12014, C5047/A8384, C5047/A15007, C5047/A10692, C8197/A16565), the National Institutes of Health (CA128978) and Post-Cancer GWAS initiative (1U19 CA148537, 1U19 CA148065 and 1U19 CA148112 - the GAME-ON initiative), the Department of Defence (W81XWH-10-1-0341), the Canadian Institutes of Health Research (CIHR) for the CIHR Team in Familial Risks of Breast Cancer, Komen Foundation for the Cure, the Breast Cancer Research Foundation, and the Ovarian Cancer Research Fund.

BPC3

The BPC3 was supported by the U.S. National Institutes of Health, National Cancer Institute (cooperative agreements U01-CA98233, U01-CA98710, U01-CA98216 and U01-CA98758 and Intramural Research Program of NIH/National Cancer Institute, Division of Cancer Epidemiology and Genetics).

CAPS

CAPS GWAS study was supported by the Cancer Risk Prediction Center (CRisP; www.crispcenter.org), a Linneus Centre (Contract ID 70867902) financed by the Swedish Research Council, (grant no K2010-70X-20430-04-3), the Swedish Cancer Foundation (grant no 09-0677), the Hedlund Foundation, the Soederberg Foundation, the Enqvist Foundation, ALF funds from the S

tockholm County Council, Stiftelsen Johanna Hagstrand och Sigfrid Linner's Minne, and Karlsson's Fund for urological and surgical research.

PEGASUS

PEGASUS was supported by the Intramural Research Program, Division of Cancer Epidemiology and Genetics, National Cancer Institute, National Institutes of Health.

ProHealth

This work was supported by National Institutes of Health grants: CA127298, CA088164, CA112355, and CA241410. This work was also supported by the UCSF Goldberg- Benioff Program in Cancer Translational Biology. Support for participant enrollment, survey completion, and biospecimen collection for the RPGEH was provided by the Robert Wood Johnson Foundation, the Wayne and Gladys Valley Foundation, the Ellison Medical Foundation, and Kaiser Permanente national and regional community benefit programs. Genotyping of the GERA cohort was funded by a grant from the National Institute on Aging, National Institute of Mental Health, and the National Institute of Health Common Fund (RC2 AG036607). We are grateful to the Kaiser Permanente Northern California members who have generously agreed to participate in the Kaiser Permanente Research Program on Genes, Environment, and Health, the ProHealth Study and the California Men's Health Study.

BBJ

The BBJ and prostate cancer GWAS in BBJ were supported by the Ministry of Education, Culture, Sports, Sciences and Technology of the Japanese government (MEXT) and the Japan Agency for Medical Research and Development (AMED).

Ghana Prostate Study (GPS)

The Ghana Prostate Study was funded by the Intramural Program of the National Cancer Institute, National Institutes of Health, Department of Health and Human Services including Contract No. HHSN261200800001E.
